# Expression of wound healing related histochemical and immunohistochemical stains at the time of salvage isolated neck dissection – an exploratory study

**DOI:** 10.64898/2026.01.11.26343877

**Authors:** Roel Henneman, Joyce Sanders, Ingrid Hofland, Olga Hamming-Vrieze, Alfons JM Balm, Erienne de Cuba

## Abstract

**Introduction:** Salvage surgery is known for an increased surgical complication rate. However, currently available patient, tumor and therapy factors are not reliable enough to predict surgical site complications (SSC). This exploratory study tests skin acquired by neck dissection for several histochemical and immunohistochemical stains to explore tissue changes related to wound healing.

**Patients and methods:** From an existing database of 232 *isolated* neck dissections (ND), seventeen patients with skin resection during ND were included - five primary and twelve salvage procedures. Skin specimens were tested for ten histochemical and immunohistochemical stains, with emphasis on the stages of wound healing; inflammation, proliferation and remodeling.

**Results:** Three stains (CD-68, α-SMA, and Vimentin), showed significant different expressions between the primary surgical and salvage groups (respectively p = 0.013, p = 0.044 and p = 0.015), without a clear pattern of decreased or increased expression. None of the ten stains showed a significant difference related to SSC.

**Conclusion:** The three significantly different staining patterns found in this exploratory study all are related to macrophage and (myo-)fibroblast transition, possibly representing an imbalance of macrophage/stroma interplay. Future studies should focus on the pivotal role of macrophages in wound healing after salvage surgery.

## Introduction

Salvage surgery of the head and neck area is known to have a higher risk of surgical site complications (SSC) as compared to primary surgery. Due to the variety in the extent of surgical procedures – with or without resection of mucosal lining and adjacent neck dissection – it is difficult to quantify this risk increasement. Neck dissections (ND) performed as an *isolated* procedure, with a clean surgical field – i.e. without contamination by saliva – are well suited for a comparison of risk. When performed as *isolated* procedure there is a limited risk of surgical site complications(1). However, performed as a salvage procedure after radiotherapy or chemoradiotherapy, the risk of serious SSCs is higher with incidences reported as high as 29%.(2)

Radiation therapy, either concurrent with chemotherapy or as sole treatment, has both early as late side effects, which can result in tissue alterations that can seriously disturb wound healing.(3, 4)

If disturbed wound healing is expected, one can try to prevent SSC by transferring well vascularized muscular tissue to the wound in order to provide ‘healthy’ non-irradiated tissue as a basis for the wound healing process, for instance by using a pedicled muscle flap intraoperatively.(5, 6) However, this means a prolonged surgical procedure with additional costs and possible donor site morbidities. Furthermore, the majority of patients will be overtreated since not each neck dissection after radiotherapy or chemoradiotherapy is complicated by SSC.

To be able to predict which patients would benefit most from the use of the transfer of well vascularized muscle tissue could therefore be helpful. Although disruption of wound healing is strongly correlated with comorbidities and a variety of other patient-dependent factors affecting the general condition, these factors are not reliable enough to predict outcome of the wound healing in an individual case.(7) Ideally, if we had preoperative knowledge about both patient factors and tissue alterations related to wound healing, it would help us to predict more accurately which patient is at high risk for disruption of wound healing. This should assist us in appropriately selecting patients for preventive use of pedicled flaps or free flap transfers.

This exploratory study was therefore intended to provide an initial insight into possible differences in tissue characteristics related to the subsequent phases of the wound healing process, in patients who underwent salvage *isolated* neck dissection with skin resection after radiotherapy or chemoradiotherapy, and with and without complication.

### Patients and methods

We searched surgical and pathology reports for skin resection during neck dissection in a well-documented database of 219 consecutive patients (232 procedures), who had undergone isolated neck dissection (as primary treatment or as salvage post-radiotherapy or post-chemoradiotherapy) at the Netherlands Cancer Institute Antoni van Leeuwenhoek from 1997 until 2012.(7) If skin resection was mentioned in one of these reports, the diagnostic HE-stained slides were reviewed.

The specimens from 17 neck-dissection procedures contained sufficient normal (i.e., non-tumorous) skin.

Informed consent was obtained of all patients before treatment, by stating that resected tissue is available for research purposes anonymously studying future options for personalized treatment. Approval of the Institutional Review Board was acquired, locally registered as CFMPB673.

The literature was searched for histochemical (HC) and immunohistochemical (IHC) staining previously used to study the characteristics of tissue involved in the different stages of healing, i.e., hemostasis, inflammation, proliferation and remodeling.

This selection process yielded the following ten stains: ALK-5A4 (anaplastic lymphoma kinase); Ki-67; CD-68; Elastica van Gieson; D2-40; alpha smooth muscle actin (α-SMA); Alcian blue; Collagen IV; CD-34; and Vimentin. Their putative involvement in the wound healing process is summarized below and in **Table 1**.

- The marker for ALK-5A4 tests for the receptor of TGF (transforming growth factor) β type 1. (8) While TGFβ 1 plays a role in normal wound healing, its expression has been found to be higher in cases of excessive fibrosis. (9) During hemostasis, TGFβ acts as a chemoattractant for monocytes, which are important to initiate the inflammation phase.(10)
- CD-68 visualizes macrophages, activated to either *M1* or *M2,* which are essential for respectively the inflammatory and the proliferative phase of wound healing. (10) And its adherence to the vascular intima and media, making it useful as a marker for vascular status. (11) CD-68 also detects fibroblasts.(12)
- Vimentin staining detects activated fibroblasts,(13) but also any monocytes and macrophages present during inflammation. We distinguished between fibroblasts and monocytes/macrophages by comparing all Vimentin-stained slides with slides stained with CD-68 (11) and hematoxylin-eosin (HE). Vimentin-deficient wounds show less fibroblast growth and prevents the initiation of epithelial-mesenchymal transition (EMT).(14)
- CD-34, a known marker for hematopoietic stem cells, is also a marker for vascular endothelial progenitors and embryonic fibroblasts. (15, 16) As result of prior trauma (e.g., radiotherapy) to the epidermis, multipotent epithelial stem cells, partly CD34 positive, migrate to the site of injury.
- D2-40 detects expression of lymphatic endothelium.(17)
- Alcian blue indicates the presence of glycosaminoglycans, an important ingredient of the extracellular matrix (ECM). (18) These affect the viscoelastic properties of the tissue by trapping water in the ECM. (16)
- As Ki-67 is used as a proliferation marker, highlighting the proliferative activity in the parabasal and basal cell layers in cases of skin damage repair. (11, 19)
- Elastica van Gieson staining indicates the presence of elastic fibers in connective tissue and also in the thickening of the vascular intima and media. (20)
- α-SMA is induced by TGFβ 1 and expressed by myofibroblasts, which are present during wound healing and are involved in wound contraction. Comparing its expression between slides helps to identify differences in myofibroblast levels in underactive and overactive healing tissue relative to normal healing tissue. (21–23)
- Collagen IV staining indicates the collagen levels present in the basal lamina and the vessels in the connective tissue. It also indicates vascular status. (24–27).

**Table 1.** Histochemical and immunohistochemical (HC/IHC) stains and their putative involvement in the wound healing process.

| Stain | Wound healing process |  |  |  |  |  |
| --- | --- | --- | --- | --- | --- | --- |
|  | Hemostasis | Inflammation | Proliferation | Remodelling | Angiogenesis | Fibrosis |
| ALK-5A4 | X |  |  |  |  | X |
| CD-68 |  | X | X |  |  | X |
| Vimentin |  | X |  |  |  | X |
| CD-34 |  |  |  |  | X | X |
| D2-40 |  |  |  |  | X |  |
| Alcian Blue |  |  |  | X |  |  |
| Ki-67 |  |  | X |  |  |  |
| Elastica von Gieson |  |  |  | X | X | X |
| α-SMA | | | X | X | | X |
| Collagen IV |  |  |  | X | X |  |

### Immunohistochemistry

Immunohistochemistry of the FFPE tumor samples was performed on a BenchMark Ultra autostainer (Ventana Medical Systems). Briefly, paraffin sections were cut at 3 μm, heated at 75°C for 28 minutes and deparaffinized in the instrument with EZ prep solution (Ventana Medical Systems). Heat-induced antigen retrieval was carried out using Cell Conditioning 1 (CC1, Ventana Medical Systems) for 8 minutes at 95°C (alpha-SMA), 32 minutes at 95°C (CD68, D2-40, CD34, Vimentin), or 64 minutes at 95°C (ALK, Ki67).

For Collagen IV enzymatic retrieval was carried out using Protease 1 (Ventana Medical Systems), 4 minutes at RT. ALK was detected using clone 5A4 (1/40 dilution, 32 minutes at 37°C, LEICA), Ki67 using clone MIB1 (1/100 dilution, 1 hour at 37°C, Agilent/DAKO), CD68 using clone KP1 (1/10.000 dilution, 32 minutes at 37°C, Agilent/DAKO), CD34 using clone QBEnd /10 (1/250 dilution, 32 minutes at 37°C, NeoMarkers), Podoplanin clone D2-40 (1/40 dilution, 32 minutes at 37°C, Agilent/DAKO), alpha-SMA using clone 1A4 (1/4000 dilution, 32 minutes at 37°C, Agilent/DAKO), Collagen IV using a cocktail (1/50 dilution, 32 minutes at 37°C, NeoMarkers) and Vimentin, clone Vim3B4 (Agilent/DAKO) at 1/5000 dilution for 32 minutes at 37 C.

For ALK and Vimentin signal amplification was applied using the Optiview Amplification Kit (Ventana Medical Systems), 4 minutes (Vimentin) or 8 minutes (ALK).

Bound antibody was detected using the OptiView DAB Detection Kit (Ventana Medical Systems). Slides were counterstained with Hematoxylin and Bluing Reagent (Ventana Medical Systems).

### Histology

Elastica van Gieson and Alcian Blue staining of the FFPE tumor samples were performed manually.

For the Elastica van Gieson staining slides were incubated with Lawson’s solution (10 minutes at 37°C, Clin-Tech Ltd.) followed by van Gieson solution (Acid Fuchsin (Bio-Optica Milano S.p.A) + Picric Acid (VWR Chemicals), 5 minutes at RT.

For Alcian Blue the slides were incubated with Alcian Blue (15 minutes at RT., Sigma-Aldrich) followed by Nuclear Fast Red (10 minutes at RT., Sigma-Aldrich). The slides were scanned with the Aperio AT2 scanner (Leica Biosystems Nussloch GmbH) and scored with the online platform Slidescore (www.slidescore.com).

### Scoring

The skin of one patient who underwent primary surgical neck dissection without surgical site complications was used as reference. Of all the other specimens, expression of the HC and IHC stains was scored on a five-point scale compared to this ND, blinded to both initial therapy and clinical outcome (JS, RH).

Results were collected together with known patient- and clinical characteristics from the existing database.

### Statistical analysis

To enable crosstab testing for the small numbers, expression outcomes were converted to a dichotomous variable. Staining patterns were divided into *normal* versus *decreased or increased.* And based on expected alteration by radiotherapy, into *reduced versus normal or increased,* respectively into *increased* versus *normal or decreased*. Testing was done by Fisher’s Exact Test or exact if the number of observations was smaller than 5 in at least one of the evaluated subgroups. P-values were two-sided or one-sided when only an increase or decrease was tested.

All statistical analyses were performed using Statistical Package for the Social Science (SPSS) version 27.0 (SPSS Inc., Chicago, IL, USA).

## Results

Pathology reports of 232 isolated neck dissections were screened. Skin tissue of sufficient quantity and quality was found in seventeen patients, 15 males and 2 females. The seventeen included neck dissections were subdivided into five primary surgeries, one post-radiotherapy and eleven post-chemoradiotherapy. Radiotherapy was given in a total dosage of 70 Gray (N = 12), concurrent chemotherapy consisted of administration of cisplatin (N = 11). The salvage procedures were grouped together. No significant difference in sex (p = 0.515) nor age (p = 0.205) was seen between the primary surgical and salvage group, see **Table 2** for all characteristics. Also, no significant difference was seen in type of ND performed (p = 0.261) or whether the sternocleidomastoid muscle was resected (p = 0.515). Insertion of a muscle flap (in all cases the pedicled myofascial pectoralis major flap) was done in 20% (1/5) in the primary surgical group and in 67% (8/12) in the salvage group (p = 0.131). Although length of surgery ranged from 48 to 350 minutes, no significant difference in median surgery time was seen between groups (p = 0.598).

**Table 2.** Patient, tumor and therapy characteristics.

| Characteristics |  |  |  |  |
| --- | --- | --- | --- | --- |
|  | Total | Primary surgery | Salvage surgery | P value |
| **Patients** |  |  |  |  |
| Number of patients | 17 | 5 | 12 |  |
| Sex (male/female) | 15/ 2 | 4/ 1 | 11/ 1 | 0.515^ |
| Age at surgery (year)<br>median (range) | 54 (28 - 83) | 66 (46-83) | 53.5 (28-65) | 0.205* |
| Tobacco consumption |  |  |  | 1.000# |
| - Never | 1 | 0 | 1 |  |
| - Former | 9 | 3 | 6 |  |
| - Active | 7 | 2 | 5 |  |
| Alcohol consumption |  |  |  | 1.000# |
| - 0-2 IU/daily | 6 | 2 | 4 |  |
| - 3-6 IU/daily | 2 | 0 | 2 |  |
| - Abuse | 9 | 3 | 6 |  |
| ASA |  |  |  | 1.000# |
| - 1 | 7 | 2 | 5 |  |
| - 2 | 9 | 3 | 6 |  |
| - 3 | 1 | 0 | 1 |  |
| BMI |  |  |  | 0.381# |
| - Underweight | 2 | 0 | 2 |  |
| - Normal weight | 11 | 3 | 8 |  |
| - Overweight | 4 | 2 | 2 |  |
| **Tumor** |  |  |  |  |
| Primary tumor |  |  |  | 0.195# |
| - Oral cavity | 3 | 1 | 2 |  |
| - Oropharynx | 10 | 2 | 8 |  |
| - Hypopharynx | 2 | 0 | 2 |  |
| - Larynx | 1 | 1 | 0 |  |
| - Other | 1 | 1 | 0 |  |
| Nodal stage |  |  |  | 1.000# |
| - N1 | 2 | 1 | 1 |  |
| - N2 | 7 | 1 | 6 |  |
| - N3 | 8 | 3 | 5 |  |
| **Therapy** |  |  |  |  |
| Type of ND |  |  |  | 0.261^ |
| - (Super) selective | 4 | 0 | 4 |  |
| - (Modified) radical | 13 | 5 | 8 |  |
| Resection SCM muscle |  |  |  | 0.515^ |
| - No | 3 | 0 | 3 |  |
| - Yes | 14 | 5 | 9 |  |
| Extracapsular spread |  |  |  | 0.294^ |
| - No | 8 | 1 | 7 |  |
| - Yes | 9 | 4 | 5 |  |
| Primary pedicled muscle<br>flap transplantation |  |  |  | 0.131^ |
| - No | 8 | 4 | 4 |  |
| - Yes | 9 | 1 | 8 |  |
| Length of surgery (minutes)<br>- median (range) | 225 (48-350) | 225 (135-328) | 215 (48-350) | 0.598* |
| ASA = American Society of Anesthesiologist scale \| BMI = body mass index \| ND = neck dissection \| SCM = sternocleidomastoid muscle |  |  |  |  |
| ^ = Fisher's Exact Test \| # = Linear by linear test \| * = Mann Whitney U test |  |  |  |  |

Seven patients (41%) experienced a surgical site complication (SSC) which required active therapy (i.e. Clavien-Dindo (CD) >1) (see **Table 3**). Wound infections were divided into superficial incisional and deep incisional, according to the Common Centers for Disease Control definitions of surgical site infection (SSI). Seven NDs were complicated by SSI, not significantly varying between both groups (p = 1.00).

**Table 3.**
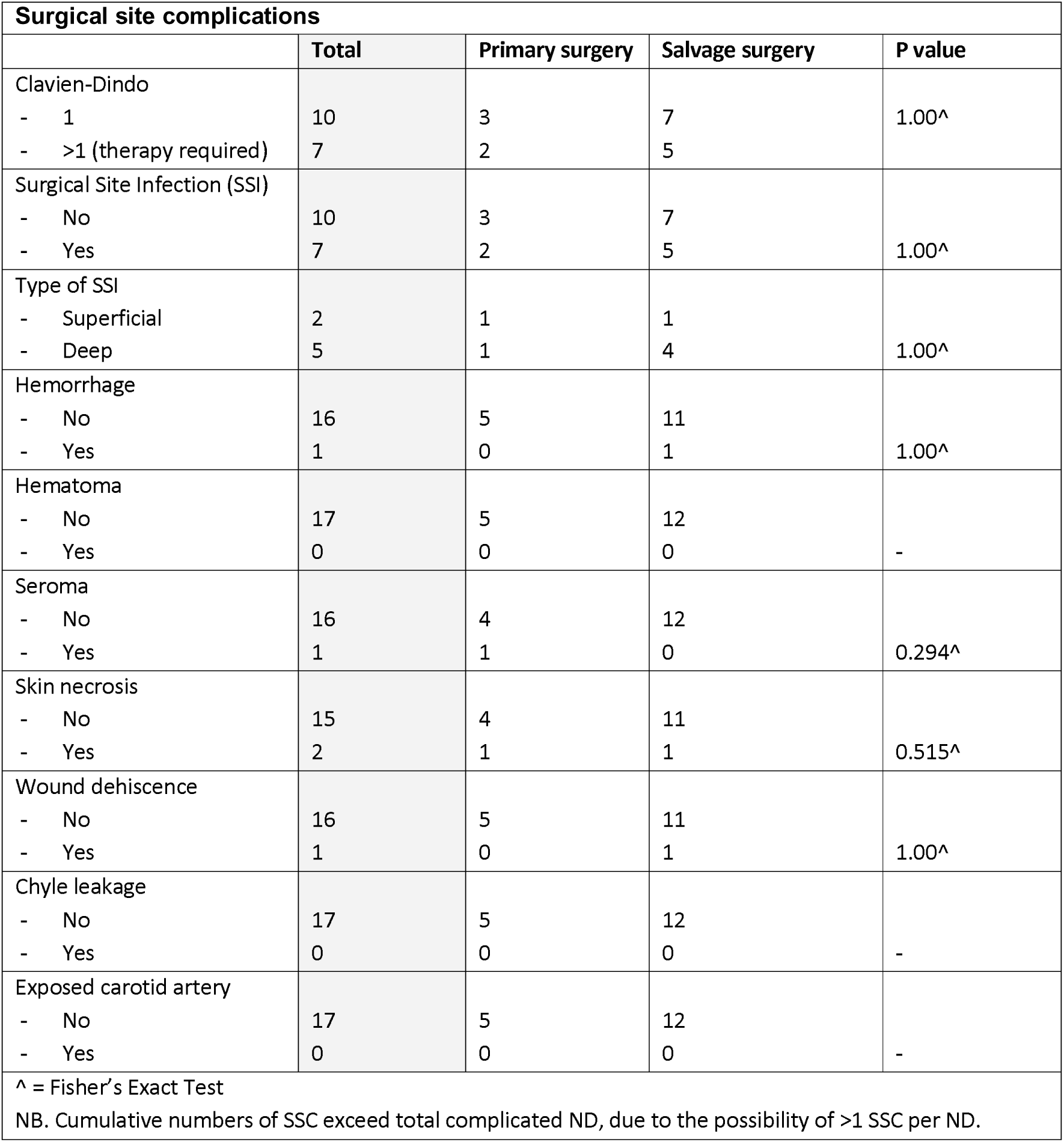
Surgical site complications.

In the primary ND group one patient developed a seroma and one had necrosis of the skin around the incision. In the salvage group, in one patient a hemorrhage led to operative intervention (CD 3b). In addition, one patient had skin necrosis and one patient had wound dehiscence.

### Histopathology

Difference in degree of fibrosis between the primary surgical and salvage group was not found by use of ALK-5A4 staining. Both in the epithelium and in the stroma no differences were seen (respectively p = 0.294 and p = 1.00). Overview of HC/IHC staining in both the primary surgical and salvage group is shown in **Table 4**.

**Table 4.**
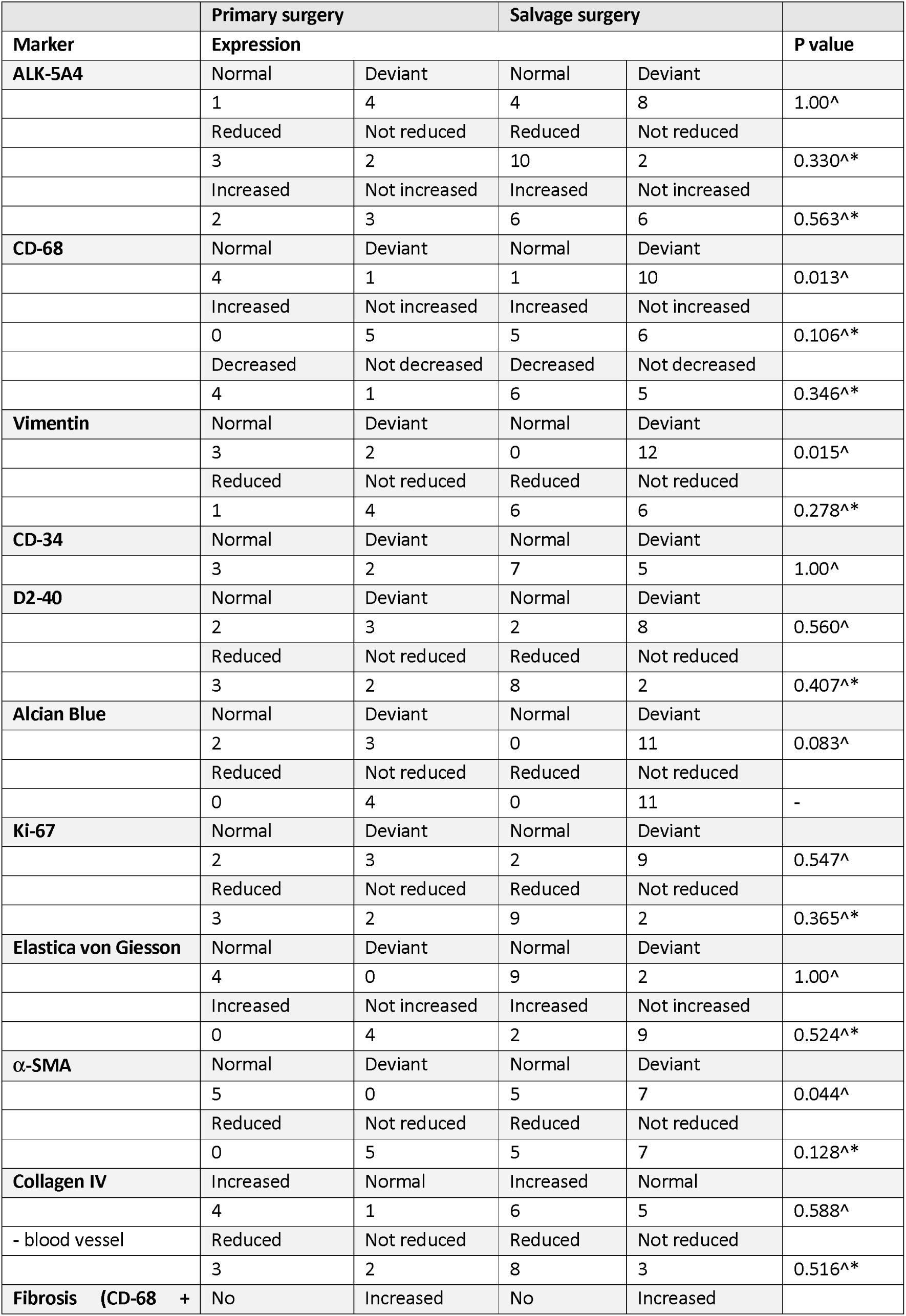
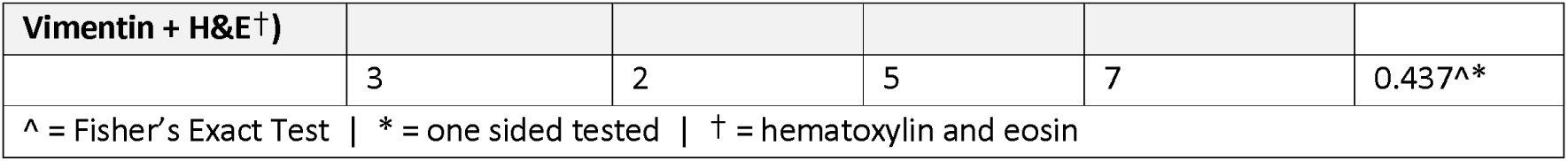
Immunohistochemical (IHC) staining patterns between surgical groups.

Expression of CD-68 in the dermis differed between these groups, p = 0.013. However, although the staining was abnormal after radiotherapy, the scoring showed either under- or overexpression without a clear trend toward either value (p = 0.106).

Also, vimentin expression was abnormal in the salvage group as compared to the control group (p = 0.015), with both under- and overexpression.

When combining expression of HE, CD-68 and Vimentin to evaluate the quantity of fibroblasts, no significant differences were found.

There were no differences observed in the quantity of blood vessels and fibroblasts (highlighted by CD-34) and lymphatic structures (highlighted by D2-40) (p = 1.00 and p = 0.560 respectively).

Alcian blue demonstrated varying staining patterns with no obvious normal pattern within both groups (p = 0.083).

The rate of proliferation by use of Ki-67 for also epithelium and stroma did not differ, p = 0.547 and p = 1.00. Elastica von Giesson staining demonstrated no difference between groups (p = 1.00); in only two salvage patients it was increased while the other 15 patients showed normal expression.

Testing α-SMA for smooth muscle cells and activated fibroblasts showed a significant difference in staining in the salvage group (p = 0.044). However, no significantly decrease nor increase in α-SMA level was seen between both surgical groups (p = 0.128).

Collagen IV staining in both stroma and as instrument to assess vascularization, showed no difference between the groups (p = 0.588 and respectively p = 0.516).

**Table 5** shows the HC and IHC staining patterns grouped by the occurrence of surgical site complication (i.e. CD 1 versus CD grade >1). For all different stains used, no significant different expressions were seen.

**Table 5.** Histochemical and immunohistochemical (HC/IHC) staining patterns for surgical site complication defined by Clavien-Dindo grade 1 and >1.

|  | Clavien-Dindo grade 1 |  | Clavien-Dindo grade >1 |  |  |
| --- | --- | --- | --- | --- | --- |
| Marker | Expression |  |  |  | P value |
| ALK-5A4 | Normal | Deviant | Normal | Deviant |  |
|  | 4 | 6 | 1 | 6 | 0.338^ |
|  | Increased | Not increased | Increased | Not increased |  |
|  | 6 | 4 | 2 | 5 | 0.218^* |
| Ki-67 | Normal | Deviant | Normal | Deviant |  |
|  | 2 | 7 | 2 | 5 | 1.00^ |
|  | Reduced | Not reduced | Reduced | Not reduced |  |
|  | 7 | 2 | 5 | 2 | 0.608^* |
| CD-68 | Normal | Deviant | Normal | Deviant |  |
|  | 4 | 5 | 1 | 6 | 0.308^ |
|  | Increased | Not increased | Increased | Not increased |  |
|  | 2 | 7 | 3 | 4 | 0.365^* |
| Elastica von Giesson | Normal | Deviant | Normal | Deviant |  |
|  | 8 | 1 | 5 | 1 | 1.00^ |
|  | Increased | Not increased | Increased | Not increased |  |
|  | 1 | 8 | 1 | 5 | 0.657^* |
| D2-40 | Normal | Deviant | Normal | Deviant |  |
|  | 2 | 7 | 2 | 4 | 1.00^ |
|  | Reduced | Not reduced | Reduced | Not reduced |  |
|  | 7 | 2 | 4 | 2 | 0.538^* |
| α-SMA | Normal | Deviant | Normal | Deviant |  |
|  | 7 | 3 | 3 | 4 | 0.350^ |
|  | Reduced | Not reduced | Reduced | Not reduced |  |
|  | 3 | 7 | 2 | 5 | 0.686^* |
| Alcian Blue | Normal | Deviant | Normal | Deviant |  |
|  | 2 | 8 | 0 | 6 | 0.500^ |
|  | Reduced | Not reduced | Reduced | Not reduced |  |
|  | 0 | 10 | 0 | 6 | - |
| Collagen IV | Increased | Normal | Increased | Normal |  |
|  | 4 | 6 | 2 | 4 | 0.608^* |
| - blood vessel | Reduced | Not reduced | Reduced | Not reduced |  |
|  | 7 | 3 | 4 | 2 | 0.654^* |
| CD-34 | Normal | Deviant | Normal | Deviant |  |
|  | 7 | 3 | 3 | 4 | 0.350^ |
| Vimentin | Normal | Deviant | Normal | Deviant |  |
|  | 1 | 9 | 5 | 2 | 0.537^ |
|  | Reduced | Not reduced | Reduced | Not reduced |  |
|  | 5 | 5 | 2 | 5 | 0.354^* |
| Fibrosis (CD-68 + Vimentin + H&E<sup>†</sup>) | No | Increased | No | Increased |  |
|  | 5 | 5 | 3 | 4 | 0.581^* |
| ^ = Fisher's Exact Test \| * = one sided tested \| † = hematoxylin and eosin |  |  |  |  |  |

^ = Fisher's Exact Test | \* = one sided tested | † = hematoxylin and eosin

## Discussion

This study was set up to explore differences in staining patterns related to the process of wound healing, which can be influenced by previous radiotherapy or chemoradiotherapy. It is a well-known clinical observation that the irradiated skin shows more fibrosis and is less well vascularized. Though, for both characteristics, there is no simple histologic grading system found.(4) Unfortunately, in this study, we were not able to unravel any unequivocal tissue alterations or immunohistochemical expression profiles, that directly relate to the impairment of normal wound healing.

Macrophages play a key-role in the normal wound healing process.(10, 28) In literature, studies report about the macrophage-to-myofibroblast transition, identified by respectively CD-68 and α-SMA,(29, 30) with vimentin acting as a mediator.(31) A low level of α-SMA could reflect less myofibroblasts, resulting in less collagen and thereby decreased ability for wound contraction.(13) A loss of vimentin finally leads to a deficient growth of fibroblasts.(14)

When comparing the specimens from the salvage surgery group to the primary surgery group significant differences are seen in CD-68, α-SMA and Vimentin expressions (respectively p = 0.013, p = 0.044 and p = 0.015), without a clear increase or decrease of staining activity. These findings suggest an imbalance in the tissue microenvironment of the radiated patients at the start of wound healing mediated by macrophages, which could potentially lead to a prolonged inflammatory phase (with M1 macrophages) without reaching next cascades in wound healing.

Also, NDs with a CDC >1 did not show a different number of macrophages than the ones with CDC 1 which might indicate that quantity counts less than the condition of individual cells (**Table 5**).

Apart from the supposed disruption of the macrophages-to-myofibrobast transition, differences in the ability of regeneration of blood vessels after primary radiotherapy could be the cause of decreased wound healing capacity as well. Although collagen IV expression appears to give more insight into vascularization (both in density of vessels and thickness of basal lamina), we were not able to make any significant correlation between the levels of expression and complications.

An attempt of combining expressions of both CD-68 and vimentin in relation to HE to distinguish from inflammation and to determine a microscopically degree of fibrosis, did not yet lead to a significant difference in both the salvage group as the group with CDC >1.

### Limitations

As this is an exploratory study, limitations are found in various aspects.

The main limiting factor for patient selection in this study was the limited availability of non-tumorous skin tissue. Skin resection is not common practice during neck dissection and is indicated in cases of suspicious invasion of tumor into the skin, thereby biasing the available skin specimens. Our interest was in tumor negative skin. However, tumor negative skin in salvage surgery specimens is scarce in pathology archives. A future prospective study could easily overcome this limitation.

Secondly, an important limitation seems to be the static aspect of this study. Skin was resected during surgery, which is the very start of the dynamic wound healing process.

The small numbers made it statistically challenging to find one-way correlations (i.e. either decrease or increase in expression) with radiotherapy and chemoradiotherapy. Also, the small numbers, prevented us to find predictive factors of wound healing problems with a bias in mind that in the salvage group a high percentage (67%) of pedicled muscle flaps were used to prevent wound healing disorders.

## Conclusion

In conclusion, significantly different expressions of α-SMA, CD-68 and Vimentin were observed in the salvage surgery group.

All these stains focus on macrophage and (myo-)fibroblast transition. The different expressions possibly represent an imbalance in the tissue microenvironment that affects macrophage function.(28) Not fully understood yet, it is likely to be caused by previous radiotherapy or chemoradiotherapy.

This finding supports the idea to conduct future studies on salvage surgery with a focus at the pivotal role of macrophages in wound healing.

### Future perspectives

Despite challenges faced, this exploratory study on HC/IHC markers – selected both on cell biology knowledge and literature – has shed some light on the complex matter of radiotherapy and chemoradiotherapy associated wound healing problem. Prediction of disturbed wound healing after salvage neck dissection remains difficult, but deserves further study. Based on our current, albeit limited, data we would propose a prospective larger study with assessment of the baseline presence of M1 and the transition to M2 macrophages and activation of fibroblasts together with the degree of vascularization at baseline.

Using routine, maybe even repeated, skin punch biopsies in the high-dose radiation fields would enable to collect larger series to study preoperatively whether patients are at risk for developing wound healing problems. These patients could take advantage of a prophylactic pedicled myofascial or free muscle flap transfer.

## Data Availability

All data produced in the present study are available upon reasonable request to the authors.

## Acknowledgements

We would like to acknowledge professor A. Sonnenberg, head of the Division of Cell Biology of the Netherlands Cancer Institute, for his help by selecting the markers. Also, we would like to acknowledge the NKI-AVL Core Facility Molecular Pathology & Biobanking (CFMPB) for supplying NKI-AVL Biobank material and lab support. And we also thank professor MWM van den Brekel and professor LE Smeele for their valuable comments.

